# Ketamine treatment effects on DNA methylation and Epigenetic Biomarkers of aging

**DOI:** 10.1101/2024.09.10.24313258

**Authors:** Kristin Dawson, Athena May Jean M. Carangan, Jessica Klunder, Natalia Carreras-Gallo, Raghav Sehgal, Samantha Megilligan, Benjamin C. Askins, Nicole Perkins, Tavis L. Mendez, Ryan Smith, Matthew Dawson, Michael Mallin, Albert T. Higgins-Chen, Varun B. Dwaraka

## Abstract

Major depressive disorder (MDD) and posttraumatic stress disorder (PTSD) are debilitating psychiatric conditions associated with poor health outcomes similarly observed in non-pathological aging. Ketamine is a dissociative anesthetic and NMDA receptor antagonist with demonstrated rapid reduction in symptoms associated with Treatment Resistant Depression (TRD) and PTSD. Ketamine’s effects on biological aging have not been extensively studied among patients with moderate to severe symptoms of depression and/or trauma. To address this gap, this study looked at the changes in non-epigenetic measures, DNA methylation levels, immune cell composition, and biological age based on various epigenetic biomarkers of aging, of 20 participants at baseline and after completion of a 2-3 week treatment course of 0.5 mg/kg ketamine infusions in individuals with MDD or PTSD. As expected, depression and PTSD scores decreased in participants following ketamine infusion treatments as measured by the PHQ-9 and PCL-5. We observed a reduction in epigenetic age in the OMICmAge, GrimAge V2, and PhenoAge biomarkers. In order to better understand the changes in epigenetic age, we also looked at the underlying levels of various Epigenetic Biomarker Proxies (EBPs) and surrogate protein markers and found significant changes following ketamine treatment. The results are consistent with existing literature on ketamine’s effects on different biomarkers. These results underline the ability of GrimAge V2, PhenoAge, and OMICmAge in particular, to capture signals associated with key clinical biomarkers, and add to the growing body of literature on ketamine’s epigenetic mechanisms and their effect on biological aging.

## Introduction

Major depressive disorder (MDD) is a debilitating condition characterized by changes in mood and affect persisting for at least two weeks (1). Posttraumatic stress disorder (PTSD) occurs in patients who have undergone or seen a traumatic event, and presents as re-experiencing traumatic events, and intense and disturbing thoughts or feelings in afflicted subjects (1). Studies have linked MDD with increased risk for functional impairments and premature mortality (2,3), as well as a variety of diseases, including cardiovascular, cerebrovascular, and metabolic disorders (4,5). Similar to MDD, PTSD is generally associated with reduced healthspan, lower quality of life, and higher risk of early death (6–8).

In recent years, there has been growing interest in utilizing various aging-related biomarkers, such as telomere length, immune cell proportions, and DNA methylation (DNAm) levels (9) to quantify biological changes that occur when individuals age to help explain adverse health outcomes. Epigenetic clocks are commonly utilized biomarkers of aging based on DNAm. First-generation epigenetic clocks predict chronological age (10,11). Second-generation biomarkers of aging, such as the GrimAge, OMICmAge, and DNAmPhenoAge, instead measure clinical features associated with aging (12,13). OMICmAge, in particular, has shown strong associations with mortality and various chronic diseases, including depression (14). Meanwhile, SystemsAge markers capture aging in 11 distinct physiological systems: blood, brain, heart, hormone, immune, inflammatory, kidney, liver, lung, metabolic, and musculoskeletal (15). Finally, the third-generation biomarker of aging, DunedinPACE, predicts the rate of aging, rather than approximating biological age (16). DNA methylation is not limited to aging clocks; it can also be used to predict metabolites, clinical measures, and proteins (14,17), often demonstrating stronger associations with clinical outcomes and disease risk than their non-DNA methylation counterparts (18,19).

Previous research has used these biomarkers to investigate the impact of MDD and PTSD on biological aging. MDD has been discovered to be linked to multiple biomarkers of DNAm age acceleration (20), as well as advanced DNAm age in blood (21). Epigenome-wide association studies have also demonstrated the involvement of differentially methylated genes in MDD in a variety of age-related biological processes, including metabolism and inflammatory response (22–24). Additionally, individuals with MDD have been found to have higher transcriptomic age compared to healthy controls (25). Likewise, PTSD is positively associated with epigenome-wide DNAm markers of increased cellular age, potentially mediated by single nucleotide polymorphisms (SNPs) in the Klotho longevity gene (26). It is also reportedly linked with an enhanced pace of epigenetic aging over time, based on measurements from the Horvath and GrimAge clocks (27). Studies have also found advanced DNAm age in the motor cortex of individuals with PTSD (28), as well as higher RNA age predictions compared to controls (29).

These commonalities between MDD, PTSD, and biological aging bring to light the potential utility of antidepressants and other PTSD medication in protecting against accelerated biological aging. In particular, ketamine, an N-methyl-D-aspartate (NMDA) receptor antagonist, has been found to induce marked improvement in depressive patients following intravenous administration, causing a drastic reduction in depressive symptoms after only three days of treatment (30). Further studies have evaluated the effectiveness of ketamine and its enantiomers in monotherapy and adjunctive therapy in treating MDD and treatment-resistant depression (TRD) (31–35). Research has also explained the prospective benefit of using ketamine on patients with PTSD, and its potential utility in reversing the synaptic abnormalities induced by PTSD (36–38). In light of these, there is reason to believe that ketamine, as an antidepressant, can potentially reduce biological aging. Literature has demonstrated the ability of psychiatric medications, including antidepressants, to decelerate aging and extend lifespan in model organisms (39–41). There is also evidence that antidepressant use is linked to lower predicted brain age (42), and lessens mortality and disease risk in humans (43,44). Taken together, these lines of evidence suggest that ketamine can contribute to reducing epigenetic aging in patients diagnosed with MDD and PTSD, without exacerbating already-increased biological aging in these diseases.

Nevertheless, there remains a dearth of information on the impact of ketamine on biological aging, as measured by molecular biomarkers and epigenetic aging biomarkers. Therefore, this study aims to comprehensively investigate the effect of a rapidly-acting agent, ketamine, on epigenetic aging in a cohort of patients diagnosed with MDD and/or PTSD whose symptoms did not remit with standard psychiatric treatment. The study will look into the individuals’ baseline DNA methylation levels, immune cell composition, and epigenetic age, and measure these parameters after ketamine administration. We hypothesize that a ketamine induction series will significantly reduce the patients’ epigenetic age predictions, immune cell subsets, and proxies of metabolites, clinical measures, and proteins after the treatment course. This pilot study shall also provide insight on the biological processes and molecular functions that mediate the effects of ketamine in depression, PTSD, and epigenetic aging.

## Materials and Methods

### Study participants and ketamine administration

To assess ketamine’s effect on biological age in patients with MDD and PTSD, 20 participants were recruited by Wild Health, Inc., Lexington, KY. All had moderate to severe symptoms despite prior antidepressant treatment. They underwent six subanesthetic ketamine infusions (0.5 mg/kg) over 2-3 weeks. Blood samples were collected at baseline and 10 days post-treatment, with 40 samples in total. One participant did not complete all infusions due to adverse effects. Samples were processed at LabCorp for clinical tests and TruDiagnostic Labs for DNA methylation analysis.

### Clinical Lab Measurement collection and analysis

The following panels were taken for the clinical screening laboratory blood tests: Urine Drug Screen and Urine Pregnancy Test (females), Complete blood count (CBC), Comprehensive metabolic panel-14 (CMP-14), Folate (Serum), Free T3, Free T4, Free Testosterone, Vitamin B12, TSH (Thyroid Stimulating Hormone), C-Reactive Protein-Cardiac, Vitamin D-25-Hydroxy, Apolipoprotein B, Hemoglobin A1c, Homocysteine, NMR LipoProfile+Lipids (Advanced Lipid Panel), Cortisol, and Insulin using clinically validated assays for baseline assessments; obtained by venipuncture for whole blood sampling at a LabCorp facility of the participant’s choosing. Only 16 patients were able to take a baseline and post-treatment measurement for the laboratory blood tests.

To look at the differences between the participants’ non-epigenetic measures, including clinical data from 60 laboratory tests and self-reported scores based on the PTSD Checklist for DSM-5 (PCL-5) and Patient Health Questionnaire-9 (PHQ-9) for MDD, we conducted a Wilcoxon signed-rank test for each set of scores in R. A p-value < 0.05 was used to denote significance.

### DNA methylation assessment

Peripheral whole blood samples were obtained using the lancet and capillary method, and preserved through mixing with lysis buffer. 500 ng of DNA was extracted and subjected to bisulfite conversion using the EZ DNA Methylation Kit from Zymo Research, following the manufacturer’s protocols. The converted samples were then randomly allocated to designated wells on the Infinium HumanMethylationEPIC BeadChip, and then subsequently amplified, hybridized onto the array, and stained. Samples were then washed and imaged using the Illumina iScan SQ instrument to capture raw image intensities.

DNAm data was pre-processed using the Minfi package in R (45–48). No outlier samples were identified using ENmix (49) and methylation values were normalized using the ssNoob method (50). We utilized the k-nearest neighbors algorithm to impute missing CpG values and finally adopted a 12-cell immune deconvolution method to estimate cell type proportions (51).

### DNA methylation biomarkers of aging and related measures

Using DNA methylation, we estimated the biological age of the participants by using the following epigenetic biomarkers of aging: DNAmPhenoAge (13), GrimAge (21), GrimAge v2 (52), and OMICmAge second-generation biomarkers, DNAmTL, which estimates telomere length, the third-generation DunedinPACE, and SystemsAge.

We used a custom R script available via GitHub (https://github.com/MorganLevineLab/PC-Clocks) to compute the principal component-based epigenetic clocks for the DNAmPhenoAge, GrimAge, GrimAge v2, and telomere length. For non-principal component-based DNAmPhenoAge epigenetic metrics, we used the methyAge function in the ENmix package in R. DunedinPACE was calculated using the PACEProjector function from the DunedinPACE package from GitHub (https://github.com/danbelsky/DunedinPACE). We calculated the SystemsAge using the authors’ script (15), which will be incorporated into the methylCIPHER package upon publication (https://github.com/MorganLevineLab/methylCIPHER).

We computed the epigenetic age acceleration (EAA) of these metrics through fitting a regression model between the participants’ chronological age and different epigenetic age measures. To control for potential batch effects, we also included in the regression model the first 3 principal components (PCs) calculated from control probes, following the methodology prescribed by Joyce et al (53). Wilcoxon signed-rank tests were performed using EAA values between timepoints at a significance level of p < 0.05. Due to the number of tests conducted for the statistical analysis of clinical test epigenetic clocks, EBP, and Marioni Episcore markers, a multi-test correction was conducted using the “number of independent degrees of freedom” approach previously described. Briefly, principal components of each measure were calculated using prcomp() in R, and the minimum number of PC’s which achieve at least 95% cumulative variance was used to adjust for Bonferroni correction (54,55).

We then conducted a correlation analysis between the biomarkers of aging and all non-epigenetic measures to investigate if the changes in lab values and questionnaire responses are concordant with changes in the predictions of biological age. Finally, we compared baseline and post-treatment levels of DNA methylation surrogate markers for multiple proteins, metabolites, and clinical variables.

### Epigenome-wide association analysis

We used the limma package to perform an epigenome-wide association study (EWAS), comparing genome-wide CpG methylation between pre- and post-treatment (56). Empirical Bayes regression models were adjusted for sex, age, batch, and three principal components. Moderated t-tests were used to assess for significance, and probes with an unadjusted p-value < 0.001 were considered significantly differentially methylated. While the false-discovery rate (FDR) was calculated, it was not used as a filter. LogFC values indicated hypermethylation (logFC > 0) or hypomethylation (logFC < 0) post-treatment. Enrichment analysis using rGREAT identified gene ontology (GO) terms associated with differentially methylated loci (DMLs) in promoter and enhancer regions.

## Results

This study analyzed clinical and epigenetic data from patients diagnosed with MDD and/or PTSD (n=20) before and after administration of ketamine infusions over the time-course. Of the 20 participants, 65% (n=13) had a comorbid diagnosis of MDD and PTSD, 10% (n=2) were diagnosed only with MDD, and 25% (n=5) had a diagnosis of only PTSD (**Supplementary Figure 1**). Mean baseline scores in the PCL-5 and PHQ-9 are also described. Blood samples were taken to evaluate DNA methylation levels among the patients to calculate their epigenetic age, the immune subsets, and the DNA methylation levels pre- and post-ketamine treatment. Subjects were made up of 75% (n=15) female and 25% (n=5) male participants, with a mean chronological age of 41.78 and 35.62, respectively. Table 1 details the demographic information of patients recruited in this study.

**Table 1.**
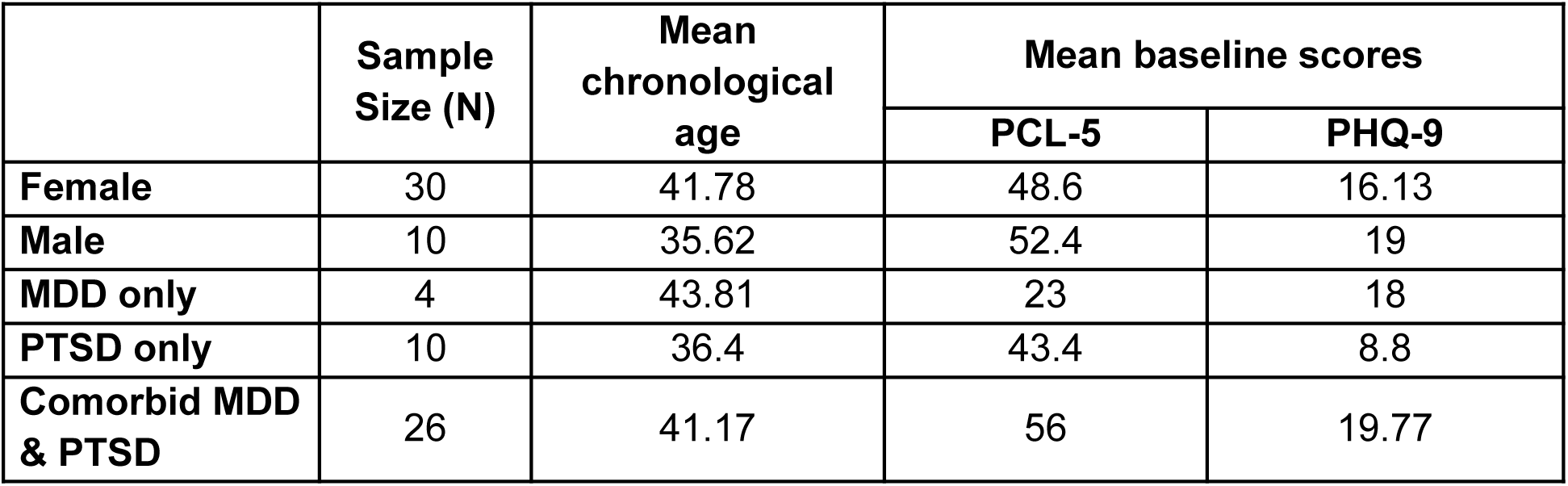
Characteristics of participants in the Ketamine trial.

|  | Sample Size (N) | Mean chronological age | Mean baseline scores |  |
| --- | --- | --- | --- | --- |
|  |  |  | PCL-5 | PHQ-9 |
| **Female** | 30 | 41.78 | 48.6 | 16.13 |
| **Male** | 10 | 35.62 | 52.4 | 19 |
| **MDD only** | 4 | 43.81 | 23 | 18 |
| **PTSD only** | 10 | 36.4 | 43.4 | 8.8 |
| **Comorbid MDD & PTSD** | 26 | 41.17 | 56 | 19.77 |

### Ketamine treatment results in a significant decline in PCL-5 and PHQ-9 scores, but not in other clinical variables

We utilized a Wilcoxon signed-rank test to compare the participant’s scores (N=20) before and after receiving the ketamine treatment course in the PCL-5 and PHQ-9. Results showed a significant decline in both PCL-5 (p=0.00000381) and PHQ-9 (p=0.0000949) scores after the participants received treatment with ketamine, with a median difference of 33 and 11 points, respectively. On the other hand, none of the laboratory tests (N=16) exhibited a significant difference following ketamine administration (**Supplementary Table 1**). Notably, a few clinical variables approached significance, such as absolute monocytes (p=0.0723), LDL-C (p=0.0743), LDL size (p=0.0829), and monocytes (p=0.0857).

### Ketamine treatment reduces epigenetic age in the third generation OMICmAge biomarker in patients with MDD and/or PTSD

Given that MDD and PTSD are associated with increased morbidity and mortality, we investigated ketamine’s effect on epigenetic biomarkers of aging most related to mortality risk. These included PhenoAge, PC PhenoAge, PC GrimAge, GrimAge v2, and OMICmAge; the DunedinPACE; and Systems Ages for blood, brain, heart, hormone, inflammation, immune, kidney, liver, lung, metabolic, musculoskeletal, and overall SystemsAge. We also investigated DNAmTL, a telomere length proxy, as telomere length has been reported to be reduced in MDD and PTSD (57). Where applicable, we utilized the principal component versions of the biomarkers, which have high test-retest reliability, which is especially important for longitudinal studies (58).

Results revealed a significant reduction in epigenetic age between timepoints, PhenoAge (p=0.024), GrimAge V2 (p=0.021), and OMICmAge (p=0.0094) (**Figure 1**). However, after adjusting by principal component variance explaining clocks, only OMICmAge retained significance (PC-adjusted Bonferonni p-value = 0.047). Other epigenetic markers did not demonstrate any significant changes; however, most epigenetic biomarkers exhibited a downward trend in epigenetic age after Ketamine treatment (**Supplementary Table 2**). Interestingly, Systems Age Inflammation approached significance, with an unadjusted p-value of 0.076.

**Figure 1.**
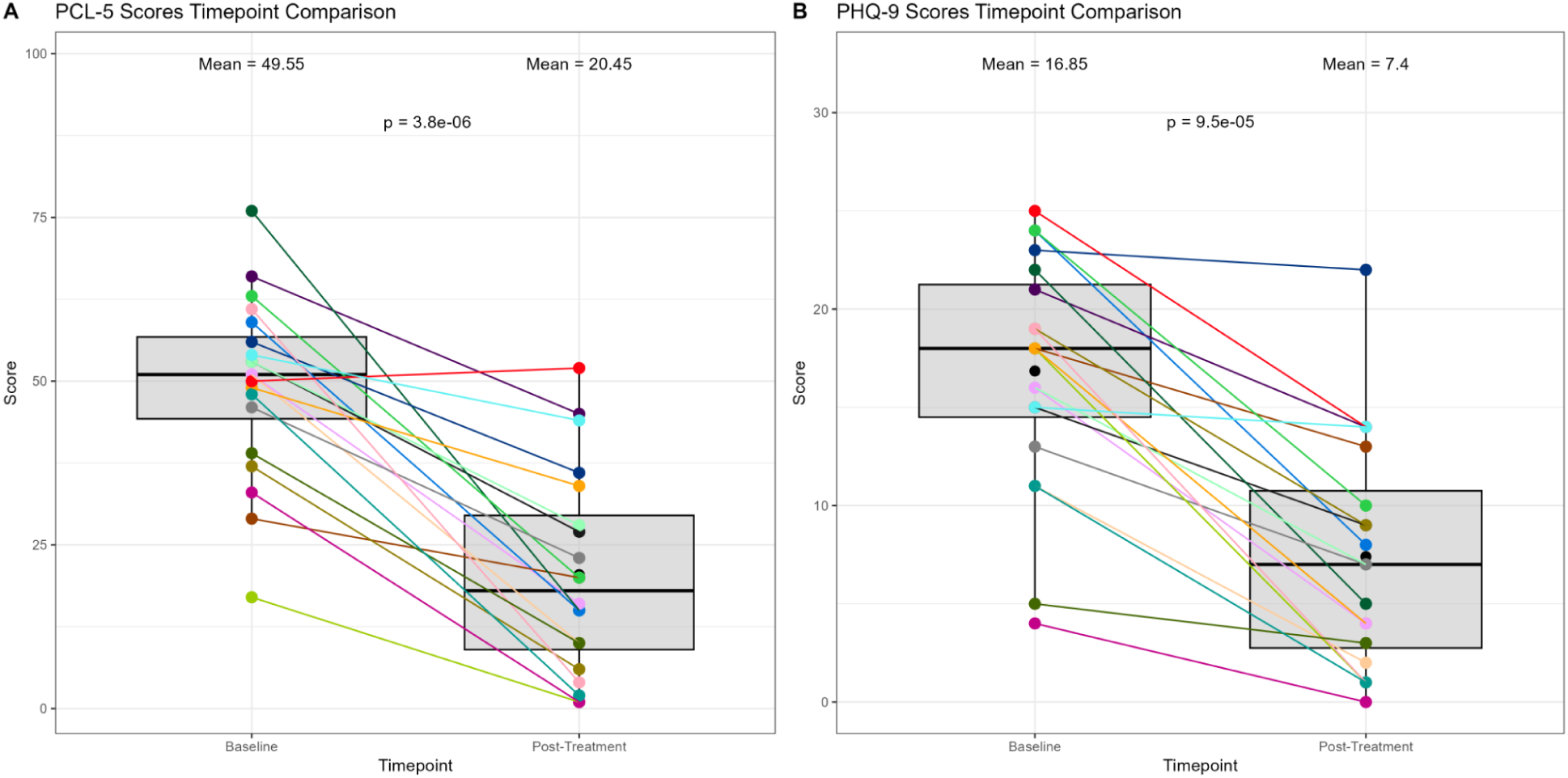
Changes in PCL-5 and PHQ-9 Scores Before and After Ketamine Treatment Course. (A) PCL-5. (B) PHQ-9. The y-axis shows the reported score for each evaluation, while the x-axis represents the timepoints for treatment with ketamine. Each color corresponds to one patient, with a line connecting their evaluation score from baseline to post-treatment.

### Ketamine treatment decreases CD4T memory cells in the blood

We quantified 12 different immune cell subsets between timepoints using EpiDISH (2023), and found a significant reduction in CD4T memory cells (unadjusted p=0.038) after ketamine treatment (**Figure 2D**). There was also a trend towards increased neutrophils (unadjusted p=0.053). Meanwhile, CD4T Naïve, CD8T Naïve, CD8T Memory, B Naïve, B Memory, Basophils, regulatory T-cells, Eosinophils, Monocytes, and Natural Killer cells did not demonstrate any notable difference after ketamine treatment (**Supplementary Table 3**).

**Figure 2.**
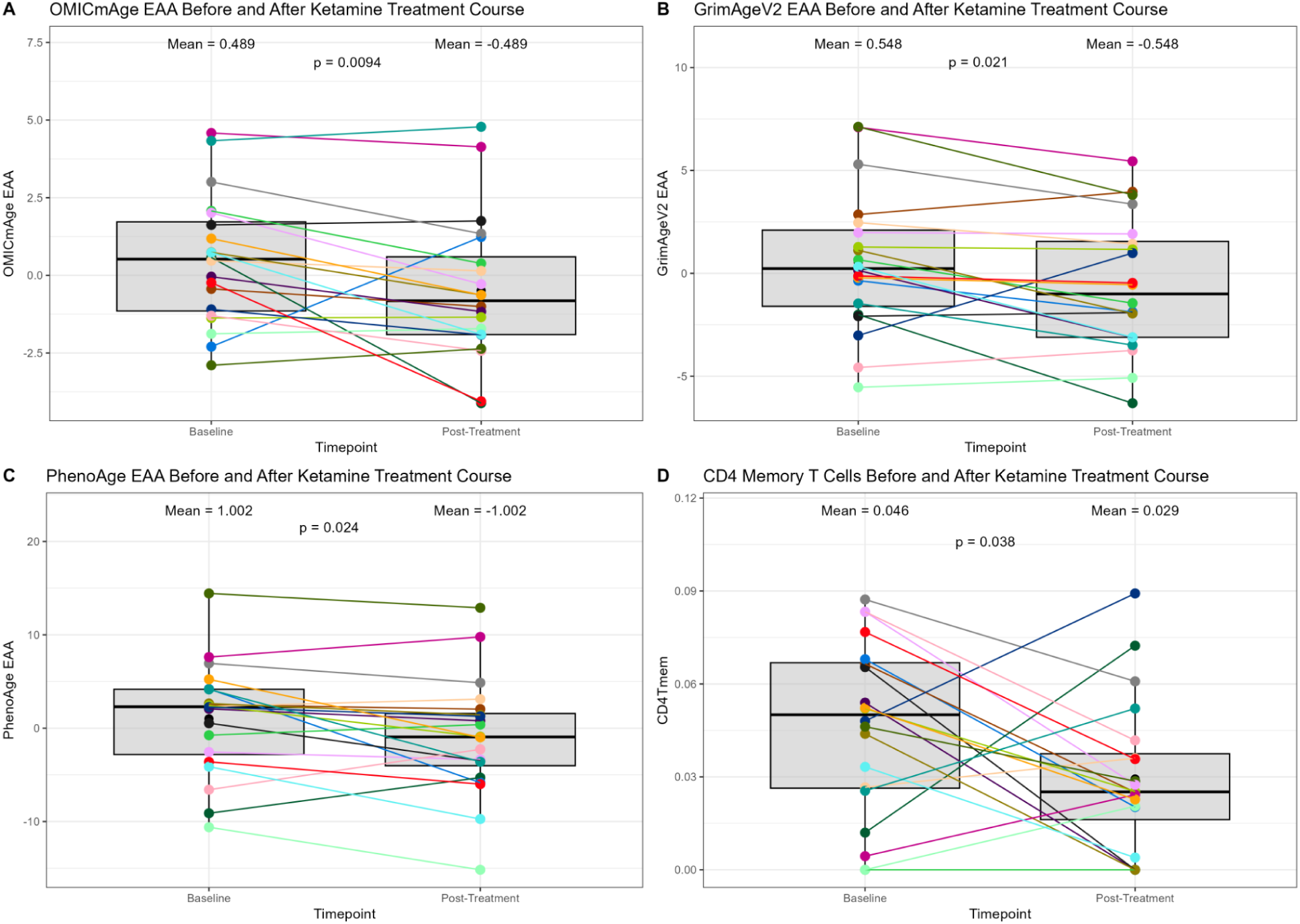
Changes in Epigenetic Biomarkers of Aging Before and After Ketamine Treatment Course. (A) OMICmAge. (B) GrimAge V2. (C) PhenoAge. (D) CD4 Memory T-cells. The y-axis shows the epigenetic age acceleration (EAA) for each biomarker of aging. The x-axis represents the timepoints for treatment with ketamine. Each color corresponds to one patient, with a line connecting their predicted EAA from baseline to post-treatment.

### Changes in clinical variables did not correlate strongly with ketamine-induced differences in epigenetic age, but were consistent among samples

To investigate whether there is a relationship between the changes in clinical variables and biological age, we conducted a correlation analysis between all these markers (**Supplementary Figure 2**). OMICmAge showed a moderate negative correlation with HbA1c (r² = -0.58, p < 0.001) and red blood cell distribution width (r² = -0.52, p < 0.001), and a positive correlation with serum folate levels (r² = 0.62, p < 0.001), which negatively correlated with PhenoAge (r² = -0.72, p < 0.001) and GrimAge V2 (r² = -0.17, p < 0.05). Both OMICmAge (r² = -0.48, p < 0.001) and GrimAge V2 (r² = -0.49, p < 0.001) showed moderate negative correlations with absolute lymphocyte count, while PhenoAge did not (r² = 0.13, p > 0.05). GrimAge V2 also had a negative correlation with lymphocyte count (r² = -0.61, p < 0.001) and a positive correlation with neutrophils (r² = 0.64, p < 0.001). PhenoAge differed by showing positive correlations with creatinine (r² = 0.52, p < 0.001), total protein (r² = 0.65, p < 0.001), WBC (r² = 0.5, p < 0.001), globulin (r² = 0.59, p < 0.001), and albumin (r² = 0.5, p < 0.001), which were weakly or negatively correlated with the other biomarkers. PhenoAge also had negative correlations with eGFR (r² = -0.58, p < 0.001) and BUN/Creatinine Ratio (r² = -0.5, p < 0.001), whereas these were positively correlated with GrimAge V2 (eGFR: r² = 0.43, p < 0.001; BUN/Creatinine Ratio: r² = 0.17, p > 0.05) and OMICmAge (eGFR: r² = 0.41, p < 0.001; BUN/Creatinine Ratio: r² = 0.33, p < 0.001). Notably, changes in PCL-5 and PHQ-9 scores did not correlate with OMICmAge, despite significant decreases in all three measures after ketamine treatment. GrimAge V2 did not significantly correlate with PCL-5 but had a weak correlation with PHQ-9 (r² = 0.04, p < 0.01). PhenoAge exhibited a negative correlation with both PCL-5 (r² = -0.39, p < 0.001) and PHQ-9 (r² = -0.16, p < 0.05).

### Ketamine has no significant effect on epigenetic biomarker proxies and Marioni protein markers

We further assessed the impact of Ketamine usage upon DNA methylation surrogate markers for multiple proteins, metabolites, and clinical variables. Results revealed 17 significantly different EBPs pre- and post-treatment (**Table 2**) out of 396 EBPs based on an PC-adjusted Bonferroni p-value < 0.05. Analysis of the Marioni Episcore protein estimates identified 13 Episcore protein markers out of 116 showed a significant (PC-adjusted Bonferroni p-value < 0.05) difference pre- and post-ketamine treatment after comparison with the Wilcox test: MMP.1 (p=0.002), NRTK3 (p=0.009), SHBG (p=0.021), Afamin (p=0.037), Contactin 4 (p=0.027), Galectin 4 (p=0.027), RARRES2 (p=0.027), and TNFRSF17 (p=0.04). However, similar to the EBPs, these markers did not reach the threshold for significance after multiple correction adjustment (False Discovery Rate, FDR), with their FDR ranging from 0.197 to 0.585 (**Table 2**).

**Table 2.** Effect of Ketamine treatment on epigenetic biomarker proxies and Marioni EpiScore markers.

| Epigenetic Biomarker Proxies |  |  |  |
| --- | --- | --- | --- |
| | Glass $\Delta$ | Wilcox $p$-value | PC-adjusted Bonferroni |
| HAP28_HUMAN | 0.01186 | 0.007296 | 0.007296 |
| Eicosenoylcarnitine (C20:1)* | 0.29262 | 0.010689 | 0.010689 |
| Glucuronide of piperine metabolite C17H21NO3 (4)* | -0.18962 | 0.015312 | 0.015312 |
| BUN_MRS | -0.02076 | 0.017181 | 0.017181 |
| adenosine | 0.21264 | 0.017181 | 0.017181 |
| malonate | 0.04982 | 0.017181 | 0.017181 |
| Ethyl alpha-glucopyranoside | 0.21809 | 0.019234 | 0.019234 |
| cytosine | 0.05052 | 0.019234 | 0.019234 |
| 3beta-hydroxy-5-cholenoate | 0.12882 | 0.023951 | 0.023951 |
| F10A1_HUMAN | -0.02942 | 0.029575 | 0.029575 |
| Eicosenedioate (C20:1-DC)* | 0.34356 | 0.029575 | 0.029575 |
| 3-methoxytyrosine | -0.0808 | 0.032768 | 0.032768 |
| deoxycarnitine | 0.21411 | 0.036234 | 0.036234 |
| BMP1_HUMAN | 0.01636 | 0.039989 | 0.039989 |
| lactose | 0.1088 | 0.044054 | 0.044054 |
| Sebacate (C10-DC) | 9.06761 | 0.048441 | 0.048441 |
| proline | -0.30485 | 0.048441 | 0.048441 |
| **Marioni EpiScores** |  |  |  |
| | Glass $\Delta$ | Wilcox p-value | PC-adjusted Bonferroni |
| NRTK3 | 0.02811 | 0.008308 | 0.008308 |
| RARRES2 | -0.05664 | 0.010689 | 0.010689 |
| MMP.1 | -0.03449 | 0.013617 | 0.013617 |
| Coagulation.factor.VII | 0.01541 | 0.019234 | 0.019234 |
| Galectin.4 | -0.02226 | 0.021484 | 0.021484 |
| Contactin.4 | 0.02742 | 0.026642 | 0.026642 |
| MMP.2 | 0.00186 | 0.029575 | 0.029575 |
| TNFRSF17 | 0.03661 | 0.029575 | 0.029575 |
| CCL10 | 0.24618 | 0.032768 | 0.032768 |
| TPO | -0.01753 | 0.032768 | 0.032768 |
| SHBG | -0.02132 | 0.036234 | 0.036234 |
| CD48.antigen | 0.00873 | 0.044054 | 0.044054 |
| Granzyme.A | 0.0149 | 0.044054 | 0.044054 |

### Ketamine treatment resulted in locus-specific epigenome wide changes

We conducted an epigenome-wide association study (EWAS) to examine DNA methylation changes after ketamine treatment, adjusting regression models for individuals, 12 immune cell levels, and principal components. To control for multiple comparisons, we used FDR and selected a model with a lambda value of 1.14, indicating no overfitting. We identified 1,144 CpG sites (p < 0.001), with 764 hypermethylated and 380 hypomethylated after treatment (**Figure 3**). Only one CpG site at Chromosome 12 (cg03703650) was significantly hypomethylated (FDR = 0.0025), located in the promoter of the advillin gene, involved in actin bundling and neurite outgrowth.

**Figure 3.**
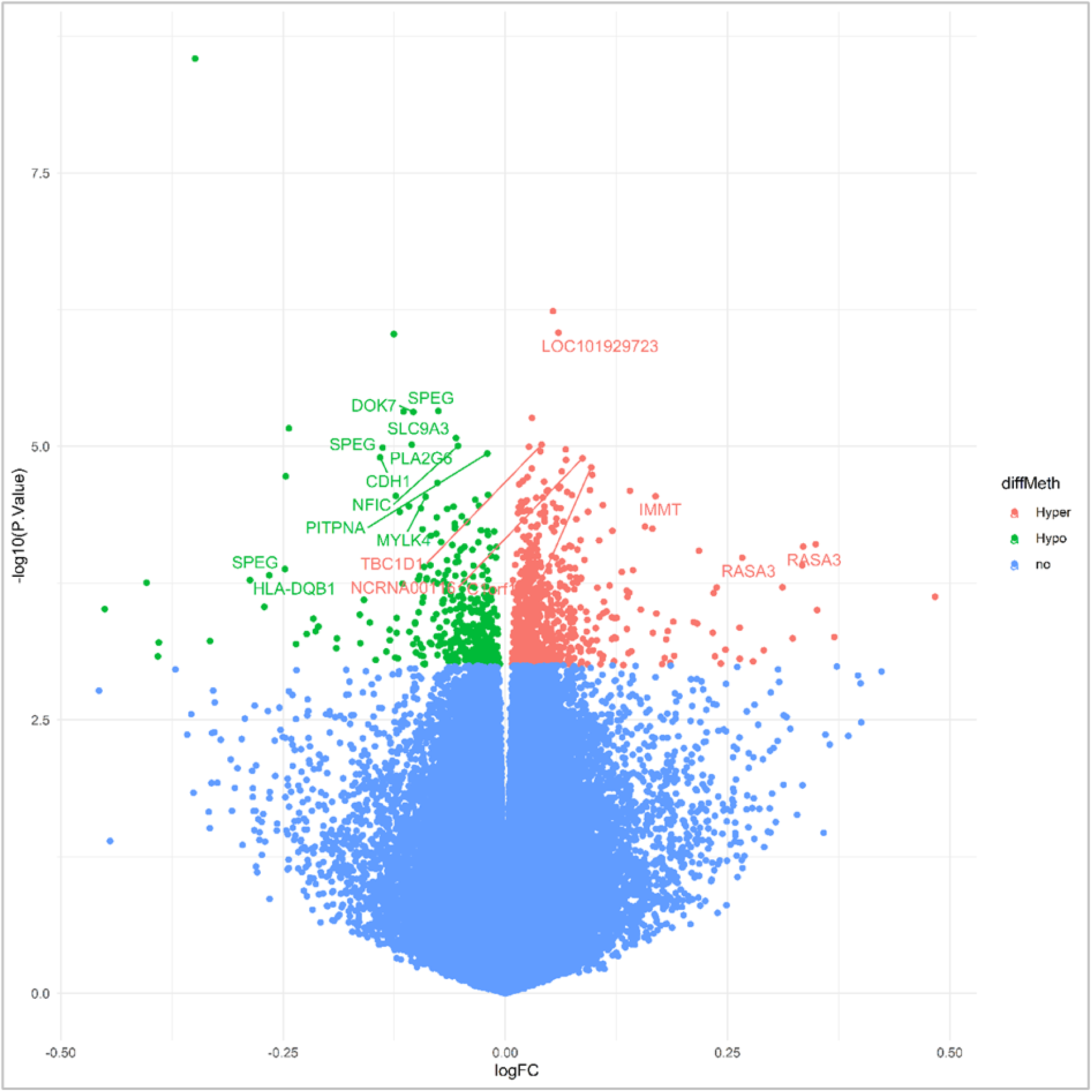
Volcano plot showing hyper- and hypomethylation of CpG sites between timepoints. The X-axis represents the log 2-fold change in DNA methylation levels between cases and controls, while the Y-axis displays the -log10 of the p-value for each CpG site. CpG sites with an FDR below 0.05 and exhibiting hypermethylation are shown in red, while those with an FDR below 0.05 and displaying hypomethylation are colored green. Moreover, we annotated the significant CpG sites with the genes where these probes are mapped. Dots without gene annotations are intergenic CpG sites.

Using the 1,144 CpG sites meeting the uncorrected p-value < 0.001, we performed two enrichment analyses, one for hypermethylated and another for hypomethylated sites. Among hypermethylated CpG sites, we identified an enrichment in processes regulating the nuclear cycle and DNA replication biological processes related to the immune system, such as regulation of T cell differentiation and mast cells, and molecular functions linked mainly to 1-phosphatidylinositol regulator activity. On the other hand, hypomethylated CpG sites were enriched in the regulation of circadian sleep, the growth plate cartilage morphogenesis, cervix development, and hindbrain tangential cell migration. We also found that these sites are mainly related to indoleamine 2,3-dioxygenase activity. The full list of associated gene ontology terms can be found in **Supplementary Table 4**.

## Discussion

This study found that administering ketamine, a rapid-acting antidepressant and off-label drug for PTSD, led to significant reductions in self-reported PTSD and MDD symptoms measured using PCL-5 and PHQ-9, as well as decreases in biological age according to the OMICmAge clock, but not among any other clock assessed. OMICmAge’s significant decline highlights its unique biomarker proxies’ ability to detect associations with depression, PTSD, and potentially other brain diseases. Created by integrating 16 protein, 14 metabolite, and 10 clinical epigenetic biomarkers across 8 biological systems, OMICmAge is particularly robust in identifying factors central to biological aging (14). In fact, previous investigation showed OMICmAge having the highest odds and hazard ratios for depression compared to other aging biomarkers (14). Ketamine administration has also been linked to changes in the metabolites and proteins used in OMICmAge, such as ribitol, uridine, triglycerides, and creatinine. Animal studies in rodents found a change in ribitol and other key elements in energy metabolism, such as phosphate and propanoic acid, post-ketamine treatment (59). Creatine and phosphocreatine have also been observed to be downregulated in the hippocampus of both rats and mice after one dose of ketamine (60,61). There also seems to be a reduction in uridine and glutamine as opposed to an increase in urea concentrations in ketamine-treated rats (60,62,63). Similarly, human studies have discovered an increase in triglycerides, cholesteryl esters, and several phosphatidylcholines in the human plasma of both healthy patients and those with treatment-resistant depression (64,65). This may explain OMICmAge’s effectiveness in capturing epigenetic aging changes in patients with MDD and/or PTSD.

We observed variation in some EBPs after ketamine administration. Prior research has linked some of these EBPs, such as proline, blood urea nitrogen (BUN), carnitines, and adenosine, to depression. For instance, high proline levels may worsen depressive symptoms via the microbiota-gut-brain axis; BUN and carnitines in combination have shown protective effects against depression (66); and BUN independently has shown protective effects against depression (35,67,68). Carnitines were found to decrease in depressive patients and subjects with a history of trauma, and carnitine supplementation has been shown to improve symptoms (69–71). Adenosine, associated with both depression and ketamine treatment, plays a role in ketamine’s anti-inflammatory and antidepressant effects by regulating glutamate neurotransmission (72). In mice models of depression, ketamine upregulated the adenosine 1 receptor, leading to antidepressant and anti-anxiety effects (73). It also increased AMPAR activation, which inhibited glutamate release, suggesting that ketamine’s antidepressant effects may involve the regulation of glutamate neurotransmission (74).

Previous studies have linked significant Marioni protein markers, such as Matrix metalloproteinases (MMPs) 1 and 2, blood coagulation factor VII, sex hormone binding globulin (SHBG), and TNFRSF17, to depression risk and prognosis. MMPs regulate inflammation and cytokine processing, with elevated MMP2 levels in cerebrospinal fluid associated with depression and schizophrenia, proportional to symptom severity. However, some studies report decreased MMP2 expression in depression (75). Coagulation factor VII has also been linked to increased depression risk (76) and suicidal behavior (77), especially in aging populations. Meanwhile, SHBG has been linked to cognitive impairment in patients with comorbid schizophrenia and depression, with elevated SHBG levels associated with higher depression risk in women, particularly post-menopause (78),74). Finally, TNFRSF17 has been found to be upregulated in women with postpartum depression (80), although no link has been established with MDD or PTSD. Together, these findings show a clear link between MDD and several EBPs and Marioni protein markers, highlighting their ability to capture disease-specific changes in MDD and PTSD.

Additionally, this study revealed pathways related to hypomethylated and hypermethylated CpG sites post-ketamine treatment, offering insights into ketamine’s antidepressant and PTSD effects. Notably, regulation of T cell differentiation was enriched in hypermethylated CpG sites, aligning with studies demonstrating the involvement of ketamine in the inhibition of the differentiation and reactivation pathogenic Th17 cells, which is upregulated in patients with MDD (81,82). Ketamine’s suppression of Th17-mediated neuroinflammation via IL-16/STAT3 inhibition supports its role in reducing CNS inflammation, a key mechanism behind its antidepressant and PTSD effects. Additionally, hypomethylation of CpG sites linked to circadian regulation suggests ketamine may improve sleep duration and quality. Sleep disorders are a known symptom and risk factor for PTSD (83,84), and research has shown an association between MDD and PTSD and poor sleep quality, resulting in abnormal mood (85–87), and aberrant expression of circadian clock genes in depressive patients (88). Ketamine’s rapid antidepressant effects may also involve resynchronizing the central clock to light cycles, causing decreased waking, and increased sleep, REM sleep, and slow-wave activity (89,90). Indeed, previous studies suggest ketamine has a chronotherapeutic effect, improving mood by shifting patients away from an “evening” body clock (91), further emphasizing the role of epigenetic changes in ketamine’s antidepressant action and its impact on epigenetic age.

Through our methylation analysis, we also discovered one CpG site with an adjusted p-value of 0.0025 associated with the advillin gene, which has been linked to axon regeneration and reduced neuropathic pain (76). Advillin is expressed in the habenula (92), which plays a role in depression and the sustained antidepressant effects of ketamine (93). However, studies have yet to establish a link between this gene and MDD or PTSD, and thus, further investigation is needed to better understand the role of the methylation of this specific location in the context of these diseases.

This pilot study identified a significant decrease in CD4T memory cells, which typically increase with age as CD4T naïve cells decline (94). CD4T memory cells are also positively associated with biological age and multimorbidity, implying that lower biological age correlates with reduced CD4 memory T-cells (95). Reduction in CD4 memory T-cells after ketamine treatment may suggest its potential to lower biological age and improve outcomes in depression and PTSD. Notably, only SystemsAge Inflammation neared significance among epigenetic aging markers, highlighting the immune system’s role in reducing epigenetic age post-treatment. The moderate negative correlation between lymphocyte count and OMICmAge and GrimAge V2 supports this link.

Further investigation is needed to understand the correlations between OMICmAge, GrimAge V2, PhenoAge, and clinical variables. Although no significant changes in lab results were observed from baseline to post-treatment, some variables approached significance. Only PCL-5 and PHQ-9 scores significantly declined following ketamine treatment, yet they did not correlate with epigenetic aging. Correlations between epigenetic aging and other clinical variables were weak, suggesting independent changes that warrant further study.

The small sample size (n=20) may have limited the detection of significant changes in aging biomarkers. Additionally, we used a fixed ketamine dose, while clinical practice often involves titration. The absence of a control group also limits validation, especially for self-reported PCL-5 and PHQ-9 scores, which can be biased. This pilot study was exploratory and requires validation in larger cohorts.

In summary, this study discovered that a 2-3 week treatment course of six ketamine infusions reduced PTSD and MDD scores, evaluated using the PCL-5 and PHQ-9. Ketamine also reduced biological age in study participants, particularly as indicated by PhenoAge, GrimAge, and OMICmAge. Our findings on altered epigenetic biomarker proxies and Marioni protein markers support their association with depression and trauma disorders, offering insights into ketamine’s clinical and epigenetic mechanisms. Additionally, we observed a decrease in CD4T memory cells, suggesting a link between ketamine and immune cell subsets, and how these may mediate a reduction in biological age. While our study supports ketamine’s role in alleviating depressive and PTSD symptoms and its potential mechanisms involving the sleep/wake cycle and neuroinflammation, further research is needed to clarify these epigenetic alterations and their contribution to ketamine’s antidepressant effects and its impact on biological age.

## Data Availability

All data produced in the present study are available upon reasonable request to the corresponding author.

## Data Availability Statement

The data that support the findings of this study are available from the corresponding authors upon reasonable request. To request data presented here, please contact the corresponding author for a data use agreement (DUA) which will detail the terms and conditions for data usage.

## Author contributions

Design and conceptualization of the study: KD, MD, MM, RS, VBD. Sample collection and recruitment: KD, JS, SM, BCA. Data generation: TLM, Processing, and statistical analyses: AC, VBD. Scientific discussion and interpretation of results: AC, NCG, RS, ATH, VBD. Wrote the manuscript: AC, RS, ATH, RS, VBD.

## Approval

The Institutional Review Board of INSTITUTE OF REGENERATIVE AND CELLULAR MEDICINE gave ethical approval for this work. Approval number: IRCM-2022-320

## References

1. DSM Library [Internet]. [cited 2024 Apr 11]. Diagnostic and Statistical Manual of Mental Disorders. Available from: https://dsm.psychiatryonline.org/doi/book/10.1176/appi.books.9780890425787

2. Cuijpers P, Vogelzangs N, Twisk J, Kleiboer A, Li J, Penninx BW. Comprehensive meta-analysis of excess mortality in depression in the general community versus patients with specific illnesses. Am J Psychiatry. 2014 Apr;171(4):453–62.

3. Brown PJ, Rutherford BR, Yaffe K, Tandler JM, Ray JL, Pott E, et al. The Depressed Frail Phenotype: The Clinical Manifestation of Increased Biological Aging. Am J Geriatr Psychiatry. 2016 Nov;24(11):1084–94.

4. Richmond-Rakerd LS, D’Souza S, Milne BJ, Caspi A, Moffitt TE. Longitudinal Associations of Mental Disorders With Dementia: 30-Year Analysis of 1.7 Million New Zealand Citizens. JAMA Psychiatry. 2022 Apr 1;79(4):333–40.

5. Leung YW, Flora DB, Gravely S, Irvine J, Carney RM, Grace SL. The impact of premorbid and postmorbid depression onset on mortality and cardiac morbidity among patients with coronary heart disease: meta-analysis. Psychosom Med. 2012 Oct;74(8):786–801.

6. Miller MW, Sadeh N. Traumatic stress, oxidative stress and post-traumatic stress disorder: neurodegeneration and the accelerated-aging hypothesis. Mol Psychiatry. 2014 Nov;19(11):1156–62.

7. Lohr JB, Palmer BW, Eidt CA, Aailaboyina S, Mausbach BT, Wolkowitz OM, et al. Is Post-Traumatic Stress Disorder Associated with Premature Senescence? A Review of the Literature. Am J Geriatr Psychiatry. 2015 Jul;23(7):709–25.

8. Wolf EJ, Logue MW, Morrison FG, Wilcox ES, Stone A, Schichman SA, et al. Posttraumatic psychopathology and the pace of the epigenetic clock: a longitudinal investigation. Psychol Med. 2019 Apr;49(5):791–800.

9. Lee E, Carreras-Gallo N, Lopez L, Turner L, Lin A, Mendez TL, et al. Exploring the effects of Dasatinib, Quercetin, and Fisetin on DNA methylation clocks: a longitudinal study on senolytic interventions. Aging. 2024 Feb 22;16(4):3088–106.

10. Horvath S. DNA methylation age of human tissues and cell types. Genome Biology. 2013 Dec 10;14(10):3156.

11. Hannum G, Guinney J, Zhao L, Zhang L, Hughes G, Sadda S, et al. Genome-wide Methylation Profiles Reveal Quantitative Views of Human Aging Rates. Molecular Cell. 2013 Jan 24;49(2):359–67.

12. Lu AT, Quach A, Wilson JG, Reiner AP, Aviv A, Raj K, et al. DNA methylation GrimAge strongly predicts lifespan and healthspan. Aging. 2019 Jan 21;11(2):303–27.

13. Levine ME, Lu AT, Quach A, Chen BH, Assimes TL, Bandinelli S, et al. An epigenetic biomarker of aging for lifespan and healthspan. Aging. 2018 Apr 18;10(4):573–91.

14. Chen Q, Dwaraka VB, Carreras-Gallo N, Mendez K, Chen Y, Begum S, et al. OMICmAge: An integrative multi-omics approach to quantify biological age with electronic medical records. bioRxiv. 2023 Oct 24;2023.10.16.562114.

15. Sehgal R, Meer M, Shadyab AH, Casanova R, Manson JE, Bhatti P, et al. Systems Age: A single blood methylation test to quantify aging heterogeneity across 11 physiological systems [Internet]. bioRxiv; 2023 [cited 2024 Jul 26]. p. 2023.07.13.548904. Available from: https://www.biorxiv.org/content/10.1101/2023.07.13.548904v1

16. Belsky DW, Caspi A, Corcoran DL, Sugden K, Poulton R, Arseneault L, et al. DunedinPACE, a DNA methylation biomarker of the pace of aging. Elife. 2022 Jan 14;11:e73420.

17. Gadd DA, Hillary RF, McCartney DL, Zaghlool SB, Stevenson AJ, Cheng Y, et al. Epigenetic scores for the circulating proteome as tools for disease prediction. eLife. 11:e71802.

18. Conole ELS, Stevenson AJ, Muñoz Maniega S, Harris SE, Green C, Valdés Hernández M del C, et al. DNA Methylation and Protein Markers of Chronic Inflammation and Their Associations With Brain and Cognitive Aging. Neurology. 2021 Dec 7;97(23):e2340–52.

19. Hillary RF, Ng HK, McCartney DL, Elliott HR, Walker RM, Campbell A, et al. Blood-based epigenome-wide analyses of chronic low-grade inflammation across diverse population cohorts. Cell Genomics [Internet]. 2024 May 8 [cited 2024 Aug 27];4(5). Available from: https://www.cell.com/cell-genomics/abstract/S2666-979X(24)00100-9

20. Liu C, Wang Z, Hui Q, Goldberg J, Smith NL, Shah AJ, et al. Association Between Depression and Epigenetic Age Acceleration: A Co-Twin Control Study. Depression and anxiety. 2022 Dec;39(12):741.

21. Protsenko E, Yang R, Nier B, Reus V, Hammamieh R, Rampersaud R, et al. “GrimAge,” an epigenetic predictor of mortality, is accelerated in major depressive disorder. Transl Psychiatry. 2021 Apr 6;11(1):1–9.

22. Zhu Y, Strachan E, Fowler E, Bacus T, Roy-Byrne P, Zhao J. Genome-wide profiling of DNA methylome and transcriptome in peripheral blood monocytes for major depression: A Monozygotic Discordant Twin Study. Transl Psychiatry. 2019 Sep 2;9(1):1–12.

23. Córdova-Palomera A, Fatjó-Vilas M, Gastó C, Navarro V, Krebs MO, Fañanás L. Genome-wide methylation study on depression: differential methylation and variable methylation in monozygotic twins. Transl Psychiatry. 2015 Apr 28;5(4):e557.

24. McKinney BC, Lin CW, Rahman T, Oh H, Lewis DA, Tseng G, et al. DNA methylation in the human frontal cortex reveals a putative mechanism for age-by-disease interactions. Transl Psychiatry. 2019 Jan 29;9(1):39.

25. Cole JJ, McColl A, Shaw R, Lynall ME, Cowen PJ, de Boer P, et al. No evidence for differential gene expression in major depressive disorder PBMCs, but robust evidence of elevated biological ageing. Transl Psychiatry. 2021 Jul 22;11(1):1–11.

26. Wolf EJ, Chen CD, Zhao X, Zhou Z, Morrison FG, Daskalakis NP, et al. Klotho, PTSD, and advanced epigenetic age in cortical tissue. Neuropsychopharmacology. 2021 Mar;46(4):721–30.

27. Hawn SE, Zhao X, Miller MW, Wallander S, Govan C, Stone A, et al. PTSD and alcohol use disorders predict the pace of cellular aging. Journal of Mood & Anxiety Disorders [Internet]. 2023 Oct 1 [cited 2024 Mar 25];3. Available from: https://www.jmoodanxdisorders.org/article/S2950-0044(23)00026-3/fulltext

28. Wolf EJ, Logue MW, Zhao X, Daskalakis NP, Morrison FG, Escarfulleri S, et al. PTSD and the Klotho Longevity Gene:Evaluation of Longitudinal Effects on Inflammation via DNA Methylation. Psychoneuroendocrinology. 2020 Jul;117:104656.

29. Kuan PF, Ren X, Clouston S, Yang X, Jonas K, Kotov R, et al. PTSD is associated with accelerated transcriptional aging in World Trade Center responders. Transl Psychiatry. 2021 May 24;11:311.

30. Berman RM, Cappiello A, Anand A, Oren DA, Heninger GR, Charney DS, et al. Antidepressant effects of ketamine in depressed patients. Biol Psychiatry. 2000 Feb 15;47(4):351–4.

31. Singh JB, Fedgchin M, Daly EJ, De Boer P, Cooper K, Lim P, et al. A Double-Blind, Randomized, Placebo-Controlled, Dose-Frequency Study of Intravenous Ketamine in Patients With Treatment-Resistant Depression. Am J Psychiatry. 2016 Aug 1;173(8):816–26.

32. Canuso CM, Singh JB, Fedgchin M, Alphs L, Lane R, Lim P, et al. Efficacy and Safety of Intranasal Esketamine for the Rapid Reduction of Symptoms of Depression and Suicidality in Patients at Imminent Risk for Suicide: Results of a Double-Blind, Randomized, Placebo-Controlled Study. Am J Psychiatry. 2018 Jul 1;175(7):620–30.

33. Popova V, Daly EJ, Trivedi M, Cooper K, Lane R, Lim P, et al. Efficacy and Safety of Flexibly Dosed Esketamine Nasal Spray Combined With a Newly Initiated Oral Antidepressant in Treatment-Resistant Depression: A Randomized Double-Blind Active-Controlled Study. Am J Psychiatry. 2019 Jun 1;176(6):428–38.

34. Daly EJ, Singh JB, Fedgchin M, Cooper K, Lim P, Shelton RC, et al. Efficacy and Safety of Intranasal Esketamine Adjunctive to Oral Antidepressant Therapy in Treatment-Resistant Depression: A Randomized Clinical Trial. JAMA Psychiatry. 2018 Feb 1;75(2):139–48.

35. Leal GC, Bandeira ID, Correia-Melo FS, Telles M, Mello RP, Vieira F, et al. Intravenous arketamine for treatment-resistant depression: open-label pilot study. Eur Arch Psychiatry Clin Neurosci. 2021 Apr 1;271(3):577–82.

36. Feder A, Parides MK, Murrough JW, Perez AM, Morgan JE, Saxena S, et al. Efficacy of Intravenous Ketamine for Treatment of Chronic Posttraumatic Stress Disorder: A Randomized Clinical Trial. JAMA Psychiatry. 2014 Jun 1;71(6):681–8.

37. Feder A, Costi S, Rutter SB, Collins AB, Govindarajulu U, Jha MK, et al. A Randomized Controlled Trial of Repeated Ketamine Administration for Chronic Posttraumatic Stress Disorder. AJP. 2021 Feb;178(2):193–202.

38. Abdallah CG, Roache JD, Averill LA, Young-McCaughan S, Martini B, Gueorguieva R, et al. Repeated ketamine infusions for antidepressant-resistant PTSD: Methods of a multicenter, randomized, placebo-controlled clinical trial. Contemporary Clinical Trials. 2019 Jun 1;81:11–8.

39. Higgins-Chen AT, Boks MP, Vinkers CH, Kahn RS, Levine ME. Schizophrenia and Epigenetic Aging Biomarkers: Increased Mortality, Reduced Cancer Risk, and Unique Clozapine Effects. Biological Psychiatry. 2020;88(3):224–35.

40. Okazaki S, Numata S, Otsuka I, Horai T, Kinoshita M, Sora I, et al. Decelerated epigenetic aging associated with mood stabilizers in the blood of patients with bipolar disorder. Transl Psychiatry. 2020 May 4;10(1):1–9.

41. Ye X, Linton JM, Schork NJ, Buck LB, Petrascheck M. A pharmacological network for lifespan extension in Caenorhabditis elegans. [cited 2024 Sep 2]; Available from: https://onlinelibrary.wiley.com/doi/10.1111/acel.12163

42. Lorenzo EC, Kuchel GA, Kuo CL, Moffitt TE, Diniz BS. Major depression and the biological hallmarks of aging. Ageing Res Rev. 2023 Jan;83:101805.

43. Taipale H, Tanskanen A, Mehtälä J, Vattulainen P, Correll CU, Tiihonen J. 20-year follow-up study of physical morbidity and mortality in relationship to antipsychotic treatment in a nationwide cohort of 62,250 patients with schizophrenia (FIN20). World Psychiatry. 2020;19(1):61–8.

44. Krivoy A, Balicer RD, Feldman B, Hoshen M, Zalsman G, Weizman A, et al. Adherence to Antidepressants Is Associated With Lower Mortality: A 4-Year Population-Based Cohort Study. J Clin Psychiatry. 2016 May 25;77(5):443.

45. Aryee MJ, Jaffe AE, Corrada-Bravo H, Ladd-Acosta C, Feinberg AP, Hansen KD, et al. Minfi: a flexible and comprehensive Bioconductor package for the analysis of Infinium DNA methylation microarrays. Bioinformatics. 2014 May 15;30(10):1363–9.

46. Fortin JP, Labbe A, Lemire M, Zanke BW, Hudson TJ, Fertig EJ, et al. Functional normalization of 450k methylation array data improves replication in large cancer studies. Genome Biology. 2014 Dec 3;15(11):503.

47. Jaffe AE, Murakami P, Lee H, Leek JT, Fallin MD, Feinberg AP, et al. Bump hunting to identify differentially methylated regions in epigenetic epidemiology studies. International Journal of Epidemiology. 2012 Feb 1;41(1):200–9.

48. Triche TJ Jr, Weisenberger DJ, Van Den Berg D, Laird PW, Siegmund KD. Low-level processing of Illumina Infinium DNA Methylation BeadArrays. Nucleic Acids Research. 2013 Apr 1;41(7):e90.

49. Xu Z, Niu L, Taylor JA. The ENmix DNA methylation analysis pipeline for Illumina BeadChip and comparisons with seven other preprocessing pipelines. Clinical Epigenetics. 2021 Dec 9;13(1):216.

50. Fortin JP, Triche TJ, Hansen KD. Preprocessing, normalization and integration of the Illumina HumanMethylationEPIC array with minfi. Bioinformatics. 2017 Feb 15;33(4):558–60.

51. Luo Q, Dwaraka VB, Chen Q, Tong H, Zhu T, Seale K, et al. A meta-analysis of immune-cell fractions at high resolution reveals novel associations with common phenotypes and health outcomes. Genome Medicine. 2023 Jul 31;15(1):59.

52. Lu AT, Binder AM, Zhang J, Yan Q, Reiner AP, Cox SR, et al. DNA methylation GrimAge version 2. Aging (Albany NY). 2022 Dec 14;14(23):9484–549.

53. Joyce BT, Gao T, Zheng Y, Ma J, Hwang SJ, Liu L, et al. Epigenetic Age Acceleration Reflects Long-Term Cardiovascular Health. Circ Res. 2021 Oct;129(8):770–81.

54. Wang C, Amini H, Xu Z, Peralta AA, Yazdi MD, Qiu X, et al. Long-term exposure to ambient fine particulate components and leukocyte epigenome-wide DNA Methylation in older men: the Normative Aging Study. Environmental Health. 2023 Aug 7;22(1):54.

55. Li MX, Yeung JMY, Cherny SS, Sham PC. Evaluating the effective numbers of independent tests and significant p-value thresholds in commercial genotyping arrays and public imputation reference datasets. Hum Genet. 2012 May 1;131(5):747–56.

56. Ritchie ME, Phipson B, Wu D, Hu Y, Law CW, Shi W, et al. limma powers differential expression analyses for RNA-sequencing and microarray studies. Nucleic Acids Research. 2015 Apr 20;43(7):e47.

57. Darrow SM, Verhoeven JE, Révész D, Lindqvist D, Penninx BW, Delucchi KL, et al. The association between psychiatric disorders and telomere length: A Meta-analysis involving 14,827 persons. Psychosom Med. 2016 Sep;78(7):776–87.

58. Higgins-Chen AT, Thrush KL, Wang Y, Minteer CJ, Kuo PL, Wang M, et al. A computational solution for bolstering reliability of epigenetic clocks: implications for clinical trials and longitudinal tracking. Nat Aging. 2022 Jul;2(7):644–61.

59. Dinis-Oliveira RJ. Metabolism and metabolomics of ketamine: a toxicological approach. Forensic Sci Res. 2017 Feb 20;2(1):2–10.

60. Lian B, Xia J, Yang X, Zhou C, Gong X, Gui S, et al. Mechanisms of ketamine on mice hippocampi shown by gas chromatography–mass spectrometry-based metabolomic analysis. NeuroReport. 2018 Jun 13;29(9):704.

61. Witkin JM, Mitchell SN, Wafford KA, Carter G, Gilmour G, Li J, et al. Comparative Effects of LY3020371, a Potent and Selective Metabotropic Glutamate (mGlu) 2/3 Receptor Antagonist, and Ketamine, a Noncompetitive N-Methyl-d-Aspartate Receptor Antagonist in Rodents: Evidence Supporting the Use of mGlu2/3 Antagonists, for the Treatment of Depression. J Pharmacol Exp Ther. 2017 Apr 1;361(1):68–86.

62. Weckmann K, Deery MJ, Howard JA, Feret R, Asara JM, Dethloff F, et al. Ketamine’s Effects on the Glutamatergic and GABAergic Systems: A Proteomics and Metabolomics Study in Mice. Mol Neuropsychiatry. 2019 Mar;5(1):42–51.

63. Wen C, Zhang M, Zhang Y, Sun F, Ma J, Hu L, et al. Brain metabolomics in rats after administration of ketamine. Biomed Chromatogr. 2016 Jan;30(1):81–4.

64. Singh B, MahmoudianDehkordi S, Voort JLV, Han X, Port JD, Frye MA, et al. Metabolomic signatures of intravenous racemic ketamine associated remission in treatment-resistant depression: A pilot hypothesis generating study. Psychiatry Research. 2022 Aug 1;314:114655.

65. Moaddel R, Zanos P, Farmer CA, Kadriu B, Morris PJ, Lovett J, et al. Comparative metabolomic analysis in plasma and cerebrospinal fluid of humans and in plasma and brain of mice following antidepressant-dose ketamine administration. Transl Psychiatry. 2022 May 2;12:179.

66. Mayneris-Perxachs J, Castells-Nobau A, Arnoriaga-Rodríguez M, Martin M, Vega-Correa L de la, Zapata C, et al. Microbiota alterations in proline metabolism impact depression. Cell Metabolism. 2022 May 3;34(5):681–701.e10.

67. Peng YF, Xiang Y, Wei YS. The significance of routine biochemical markers in patients with major depressive disorder. Sci Rep. 2016 Sep 29;6:34402.

68. Mao Y, Li X, Zhu S, Ma J, Geng Y, Zhao Y. Associations between urea nitrogen and risk of depression among subjects with and without type 2 diabetes: A nationwide population-based study. Front Endocrinol [Internet]. 2022 Oct 27 [cited 2024 Jun 7];13. Available from: https://www.frontiersin.org/journals/endocrinology/articles/10.3389/fendo.2022.985167/full

69. Wang SM, Han C, Lee SJ, Patkar AA, Masand PS, Pae CU. A review of current evidence for acetyl-l-carnitine in the treatment of depression. J Psychiatr Res. 2014 Jun;53:30–7.

70. Nasca C, Bigio B, Lee FS, Young SP, Kautz MM, Albright A, et al. Acetyl-l-carnitine deficiency in patients with major depressive disorder. Proc Natl Acad Sci U S A. 2018 Aug 21;115(34):8627–32.

71. Liu T, Deng K, Xue Y, Yang R, Yang R, Gong Z, et al. Carnitine and Depression. Front Nutr. 2022 Mar 14;9:853058.

72. Mazar J, Rogachev B, Shaked G, Ziv NY, Czeiger D, Chaimovitz C, et al. Involvement of adenosine in the antiinflammatory action of ketamine. Anesthesiology. 2005 Jun;102(6):1174–81.

73. Serchov T, Clement HW, Schwarz MK, Iasevoli F, Tosh DK, Idzko M, et al. Increased Signaling via Adenosine A1 Receptors, Sleep Deprivation, Imipramine, and Ketamine Inhibit Depressive-like Behavior via Induction of Homer1a. Neuron. 2015 Aug 5;87(3):549–62.

74. Lazarevic V, Yang Y, Flais I, Svenningsson P. Ketamine decreases neuronally released glutamate via retrograde stimulation of presynaptic adenosine A1 receptors. Mol Psychiatry. 2021;26(12):7425–35.

75. Bobińska K, Szemraj J, Gałecki P, Talarowska M. The role of MMP genes in recurrent depressive disorders and cognitive functions. Acta Neuropsychiatrica. 2016 Aug;28(4):221–31.

76. Doulalas AD, Rallidis LS, Gialernios T, Moschonas DN, Kougioulis MN, Rizos I, et al. Association of depressive symptoms with coagulation factors in young healthy individuals. Atherosclerosis. 2006 May 1;186(1):121–5.

77. Yang Y, Chen J, Liu C, Fang L, Liu Z, Guo J, et al. The Extrinsic Coagulation Pathway: a Biomarker for Suicidal Behavior in Major Depressive Disorder. Sci Rep. 2016 Sep 8;6:32882.

78. Colangelo LA, Craft LL, Ouyang P, Liu K, Schreiner PJ, Michos ED, et al. Association of Sex Hormones and SHBG with Depressive Symptoms in Post-menopausal Women: the Multi-Ethnic Study of Atherosclerosis. Menopause. 2012 Aug;19(8):877–85.

79. Zhu H, Sun Y, Guo S, Zhou Q, Jiang Y, Shen Y, et al. Causal relationship between sex hormone-binding globulin and major depression: A Mendelian randomization study. Acta Psychiatrica Scandinavica. 2023;148(5):426–36.

80. Mehta D, Grewen K, Pearson B, Wani S, Wallace L, Henders AK, et al. Genome-wide gene expression changes in postpartum depression point towards an altered immune landscape. Transl Psychiatry. 2021 Mar 4;11:155.

81. Lee JE, Lee JM, Park YJ, Kim BS, Jeon YT, Chung Y. Inhibition of autoimmune Th17 cell responses by pain killer ketamine. Oncotarget. 2017 May 31;8(52):89475–85.

82. Davami MH, Baharlou R, Ahmadi Vasmehjani A, Ghanizadeh A, Keshtkar M, Dezhkam I, et al. Elevated IL-17 and TGF-β Serum Levels: A Positive Correlation between T-helper 17 Cell-Related Pro-Inflammatory Responses with Major Depressive Disorder. Basic Clin Neurosci. 2016 Apr;7(2):137–42.

83. Lewis C, Lewis K, Kitchiner N, Isaac S, Jones I, Bisson JI. Sleep disturbance in post-traumatic stress disorder (PTSD): a systematic review and meta-analysis of actigraphy studies. European Journal of Psychotraumatology. 2020 Dec 31;11(1):1767349.

84. Lancel M, van Marle HJF, Van Veen MM, van Schagen AM. Disturbed Sleep in PTSD: Thinking Beyond Nightmares. Front Psychiatry [Internet]. 2021 Nov 24 [cited 2024 Jul 18];12. Available from: https://www.frontiersin.org/journals/psychiatry/articles/10.3389/fpsyt.2021.767760/full

85. Cheng W, Rolls ET, Ruan H, Feng J. Functional Connectivities in the Brain That Mediate the Association Between Depressive Problems and Sleep Quality. JAMA Psychiatry. 2018 Oct 1;75(10):1052–61.

86. Bunney BG, Li JZ, Walsh DM, Stein R, Vawter MP, Cartagena P, et al. Circadian dysregulation of clock genes: clues to rapid treatments in major depressive disorder. Mol Psychiatry. 2015 Feb;20(1):48–55.

87. Neylan TC, Metzler TJ, Best SR, Weiss DS, Fagan JA, Liberman A, et al. Critical incident exposure and sleep quality in police officers. Psychosom Med. 2002;64(2):345–52.

88. Li JZ, Bunney BG, Meng F, Hagenauer MH, Walsh DM, Vawter MP, et al. Circadian patterns of gene expression in the human brain and disruption in major depressive disorder. Proc Natl Acad Sci U S A. 2013 Jun 11;110(24):9950–5.

89. Duncan WC, Sarasso S, Ferrarelli F, Selter J, Riedner BA, Hejazi NS, et al. Concomitant BDNF and sleep slow wave changes indicate ketamine-induced plasticity in major depressive disorder. Int J Neuropsychopharmacol. 2013 Mar;16(2):301–11.

90. Duncan WC, Ballard ED, Zarate CA. Ketamine-Induced Glutamatergic Mechanisms of Sleep and Wakefulness: Insights for Developing Novel Treatments for Disturbed Sleep and Mood. Handb Exp Pharmacol. 2019;253:337–58.

91. Yan R, Marshall T, Khullar A, Nagle T, Knowles J, Malkin M, et al. Patient-reported outcomes on sleep quality and circadian rhythm during treatment with intravenous ketamine for treatment-resistant depression. Ther Adv Psychopharmacol. 2024;14:20451253241231264.

92. Hunter DV, Smaila BD, Lopes DM, Takatoh J, Denk F, Ramer MS. Advillin Is Expressed in All Adult Neural Crest-Derived Neurons. eNeuro. 2018;5(5):ENEURO.0077-18.2018.

93. Ma S, Chen M, Jiang Y, Xiang X, Wang S, Wu Z, et al. Sustained antidepressant effect of ketamine through NMDAR trapping in the LHb. Nature. 2023 Oct;622(7984):802–9.

94. Nikolich-Zugich J. T cell aging: naive but not young. J Exp Med. 2005 Mar 21;201(6):837–40.

95. Moro-García MA, Alonso-Arias R, López-Larrea C. When Aging Reaches CD4+ T-Cells: Phenotypic and Functional Changes. Front Immunol. 2013 May 10;4:107.

