## Supplementary Materials for "Ketamine treatment effects on DNA methylation and Epigenetic Biomarkers of aging"

Supplementary Material

### Supplementary Tables

**Supplementary Table 1. Effect of ketamine treatment series on non-epigenetic variables.** Mean and median values are calculated at baseline and after administration of ketamine. The final column shows the p-value of the difference between treatments. Significantly changed terms are bolded.

| | Mean | | Median | | p-value | PC-Bonferroni |
|---|---|---|---|---|---|---|
| | Pre- | Post- | Pre- | Post- | | |
| **PCL-5** | **49.55** | **20.45** | **51** | **18** | **0.0000038** | **0.0000076** |
| **PHQ-9** | **16.85** | **7.4** | **18** | **7** | **0.0000949** | **0.0001898** |
| A/G Ratio | 1.913 | 1.969 | 1.9 | 1.9 | 0.1563 | 0.3126 |
| Albumin | 4.431 | 4.487 | 4.45 | 4.5 | 0.3253 | 0.6506 |
| Alkaline Phosphatase | 72.562 | 72.688 | 66.5 | 68 | 0.8764 | 1.000 |
| ALT (SGPT) | 19.188 | 17.875 | 18 | 16.5 | 0.1917 | 0.3834 |
| Apolipoprotein B | 82.25 | 83.188 | 83 | 82 | 0.8155 | 1.000 |
| AST (SGOT) | 19.875 | 18.875 | 18 | 18.5 | 0.0863 | 0.1726 |
| Basophils (Absolute) | 0.019 | 0.031 | 0 | 0 | 0.4237 | 0.8474 |
| Basophils | 0.75 | 0.812 | 1 | 1 | 0.7656 | 1.000 |
| Bilirubin, Total | 0.423 | 0.523 | 0.4 | 0.4 | 0.1747 | 0.3494 |
| BUN | 12.188 | 12 | 11 | 11 | 0.8243 | 1.000 |

| | | | | | | |
|---|---|---|---|---|---|---|
| BUN/Creatinine Ratio | 14 | 13.75 | 12.5 | 13 | 0.7786 | 1.000 |
| C-Reactive Protein, Cardiac | 2.492 | 3.5 | 1.195 | 0.59 | 0.2412 | 0.4824 |
| Calcium | 9.456 | 9.456 | 9.6 | 9.45 | 0.4889 | 0.9778 |
| Carbon Dioxide, Total | 23.625 | 24.688 | 23 | 24 | 0.0776 | 0.1552 |
| Chloride | 102.25 | 102.125 | 102 | 102.5 | 0.8050 | 1.000 |
| Cholesterol, Total | 181.562 | 190.625 | 184 | 180.5 | 0.1704 | 0.3408 |
| Cortisol | 12.938 | 11.325 | 13.8 | 11.65 | 0.2744 | 0.5488 |
| Creatinine | 0.869 | 0.883 | 0.835 | 0.87 | 0.5925 | 1.000 |
| eGFR | 94.625 | 92.062 | 92 | 92.5 | 0.4887 | 0.9774 |
| Eosinophils | 2.688 | 2.688 | 2 | 2 | 0.4627 | 0.9254 |
| Eosinophils (Absolute) | 0.144 | 0.169 | 0.1 | 0.1 | 1 | 2 |
| Folate (Folic Acid), Serum | 9.875 | 9.088 | 10.35 | 7.95 | 0.6726 | 1.000 |
| Free Testosterone (Direct) | 4.486 | 3.764 | 2.25 | 2.3 | 0.975 | 1.000 |
| Globulin, Total | 2.369 | 2.331 | 2.45 | 2.25 | 0.609 | 1.000 |
| Glucose | 92 | 93.125 | 93.5 | 93 | 0.5516 | 1.000 |
| HDL-C | 57.5 | 56.562 | 64.5 | 59 | 0.842 | 1.000 |
| HDL-P | 34.425 | 34.444 | 34.5 | 35.3 | 1 | 1.000 |
| Hematocrit | 41.512 | 41.219 | 41.4 | 40.3 | 0.7173 | 1.000 |
| Hemoglobin | 13.85 | 13.85 | 13.9 | 13.6 | 0.9587 | 1.000 |
| Hemoglobin A1c | 5.4 | 5.388 | 5.4 | 5.5 | 0.5885 | 1.000 |
| Homocysteine | 8.988 | 9.488 | 8.85 | 9.25 | 0.1543 | 0.3086 |

| | | | | | | |
|---|---|---|---|---|---|---|
| Immature Granulocytes | 0.071 | 0 | 0 | 0 | 1 | 1.000 |
| Insulin | 12.1 | 12.481 | 9 | 9.2 | 0.7057 | 1.000 |
| LDL-C (NIH Calc) | 104.812 | 113.688 | 99 | 105.5 | 0.0743 | 0.1486 |
| LDL-P | 1245.357 | 1258.071 | 1231 | 1175.5 | 0.4897 | 0.9794 |
| LDL Size | 21.2 | 21.4 | 21.3 | 21.4 | 0.0829 | 0.1658 |
| LP-IR Score | 59.125 | 51.25 | 57 | 52 | 0.1083 | 0.2166 |
| Lymphocytes | 34.312 | 34.688 | 34.5 | 35.5 | 0.6596 | 1.000 |
| Lymphocytes (Absolute) | 1.8 | 1.9 | 1.7 | 1.75 | 0.3301 | 0.6602 |
| MCH | 30.275 | 30.262 | 30.25 | 29.95 | 0.7365 | 1.000 |
| MCHC | 33.388 | 33.606 | 32.9 | 33.6 | 0.3254 | 0.6508 |
| MCV | 90.75 | 90.125 | 91 | 90.5 | 0.1411 | 0.2822 |
| Monocytes | 8.5 | 7.875 | 8 | 8 | 0.0857 | 0.1714 |
| Monocytes (Absolute) | 0.469 | 0.431 | 0.4 | 0.4 | 0.0723 | 0.1446 |
| Neutrophils | 53.688 | 53.938 | 52 | 53.5 | 0.8423 | 1.000 |
| Neutrophils (Absolute) | 3.138 | 3.038 | 3.05 | 2.6 | 0.7796 | 1.000 |
| Platelets | 261.312 | 273.188 | 260.5 | 277 | 0.2242 | 0.4484 |
| Potassium | 4.412 | 4.312 | 4.5 | 4.35 | 0.2038 | 0.4076 |
| Protein, Total | 6.8 | 6.819 | 6.7 | 6.8 | 0.7748 | 1.000 |
| RBC | 4.585 | 4.582 | 4.61 | 4.48 | 0.8971 | 1.000 |
| RDW | 12.656 | 12.525 | 12.7 | 12.4 | 0.2545 | 0.509 |
| Small LDL-P | 389.385 | 334 | 344 | 343 | 0.2459 | 0.4918 |
| Sodium | 140 | 139.75 | 140 | 140 | 0.7172 | 1.000 |
| T4, Free (Direct) | 1.204 | 1.22 | 1.185 | 1.245 | 0.7981 | 1.000 |

| | | | | | | |
|---|---|---|---|---|---|---|
| Triglycerides | 106.812 | 113 | 92.5 | 98.5 | 0.6292 | 1.000 |
| Triiodothyronine (T3), Free | 3.212 | 3.175 | 3.2 | 3.15 | 0.7062 | 1.000 |
| TSH | 1.879 | 1.726 | 1.88 | 1.52 | 0.5282 | 1.000 |
| Vitamin B12 | 581.938 | 581.188 | 564 | 612.5 | 0.7057 | 1.000 |
| Vitamin D, 25-Hydroxy | 37.594 | 37.225 | 34.05 | 35.35 | 0.9399 | 1.000 |
| WBC | 5.575 | 5.581 | 5.3 | 5.2 | 0.7366 | 1.000 |

**Supplementary Table 2. Impact of Ketamine treatment on epigenetic age**. The average change in epigenetic age acceleration (EAA) for 17 epigenetic clocks is shown, as measured at baseline and after administration of ketamine. The final column shows the p-value of the difference between treatments. Clocks exhibiting significant changes in their epigenetic age between timepoints are bolded.

| | **Glass Δ** | **Standard Error Difference** | **p-value** | **PCA Adj. Bonferroni** |
|---|---|---|---|---|
| PC PhenoAge EAA | 0.98 | 1.759 | 0.2 | 1.000 |
| PC DNAmTL EAA | 0.0042 | 0.028 | 0.7 | 1.000 |
| PC GrimAge EAA | -0.34 | 0.660 | 0.29 | 1.000 |
| **GrimAge V2 EAA** | **-1.1** | | **0.021** | 0.105 |
| **OMICmAge EAA** | **-0.98** | **0.693** | **0.0094** | **0.047** |
| DunedinPACE | 0.01 | | 0.87 | 1.000 |
| **PhenoAge EAA** | **-2** | **0.043** | **0.024** | 0.120 |
| Systems Age Blood EAA | -0.56 | 2.545 | 0.55 | 1.000 |
| Systems Age Brain EAA | -1.48 | 3.691 | 0.41 | 1.000 |
| Systems Age Inflammation EAA | -1.9 | 3.544 | 0.076 | 0.380 |
| Systems Age Heart EAA | -0.82 | 3.057 | 0.37 | 1.000 |
| Systems Age Hormone EAA | -0.42 | 1.613 | 0.78 | 1.000 |

| | | | | |
|---|---|---|---|---|
| Systems Age Immune EAA | -0.98 | 3.054 | 0.41 | 1.000 |
| Systems Age Kidney EAA | -1.28 | 3.069 | 0.28 | 1.000 |
| Systems Age Liver EAA | -1.38 | 2.999 | 0.33 | 1.000 |
| Systems Age Lung EAA | -1.08 | 2.503 | 0.29 | 1.000 |
| Systems Age Metabolic EAA | -2.94 | 4.070 | 0.12 | 0.600 |
| Systems Age Musculoskeletal EAA | -2.78 | 3.555 | 0.11 | 0.550 |
| SystemsAge EAA | -1.1 | 3.135 | 0.29 | 1.000 |

**Supplementary Table 3. Comparison of immune cell subsets before and after Ketamine treatment.** Mean and median values for 12 immune subsets estimated using DNA methylation are calculated at baseline and after administration of ketamine. The final column shows the p-value of the difference between timepoints. The significant differences are bolded.

| | **Mean** | | **Median** | | **p-value** |
|---|---|---|---|---|---|
| | **Pre-treatment** | **Post-treatment** | **Pre-treatment** | **Post-treatment** | |
| **CD4T Naive** | 0.083 | 0.073 | 0.084 | 0.069 | 0.18 |
| **CD4T Memory** | **0.046** | **0.029** | **0.05** | **0.025** | **0.038** |
| **CD8T Naive** | 0.029 | 0.014 | 0.013 | 0.004 | 0.17 |
| **CD8T Memory** | 0.054 | 0.062 | 0.044 | 0.052 | 0.35 |
| **B naive** | 0.033 | 0.023 | 0.033 | 0.018 | 0.12 |
| **B memory** | 0.014 | 0.013 | 0.011 | 0.013 | 0.93 |
| **Basophil** | 0.012 | 0.009 | 0.012 | 0.009 | 0.1 |
| **Regulatory T cells** | 0.033 | 0.038 | 0.03 | 0.034 | 0.19 |
| **Eosinophil** | 0.003 | 0.001 | 0 | 0 | 0.14 |
| **Natural Killer** | 0.028 | 0.023 | 0.019 | 0.025 | 0.62 |
| **Neutrophil** | 0.626 | 0.687 | 0.612 | 0.675 | 0.053 |
| **Monocyte** | 0.04 | 0.029 | 0.04 | 0.034 | 0.11 |

**Supplementary Figure 1. Scatter Plot of Individual Scores at Baseline.** The x-axis shows the treatment groups and the y-axis corresponds to the patients' self-reported score for each test at baseline. The colors correspond to the patients' diagnosis: MDD alone, PTSD alone, or comorbid MDD and PTSD. The red dashed line corresponds to the minimum score needed for a diagnosis of moderately severe depression based on the PHQ-9. The green dashed line corresponds to the minimum score needed for a diagnosis of PTSD.

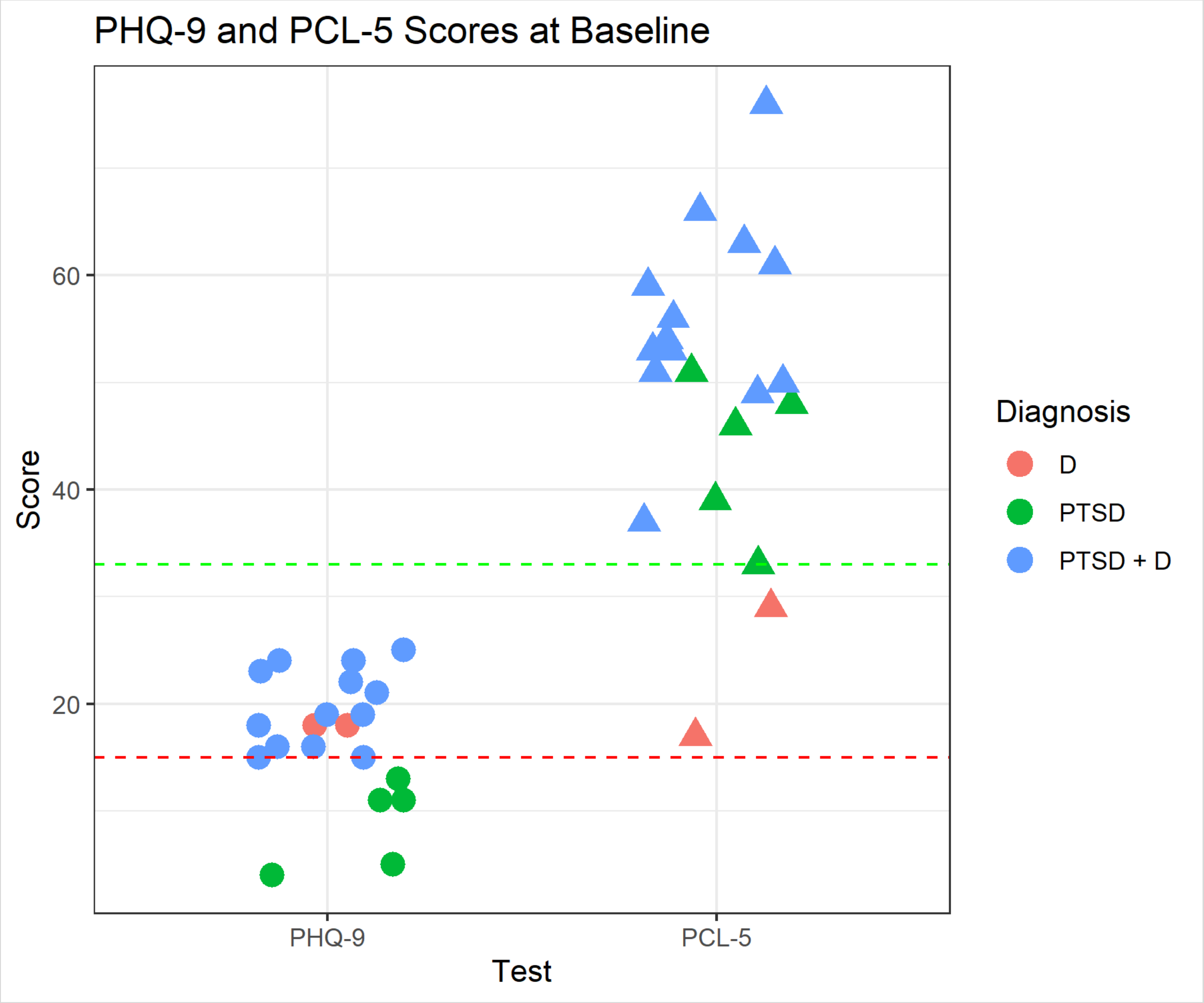


**Supplementary Figure 2. Correlation plot showing the strength of association between changes in non-epigenetic measures and epigenetic age from baseline to post-treatment.** This correlation plot has all variables of interest in the x- and y-axis. The strength of correlation is denoted by the size and color of the circles in each cell. Coefficient values are listed in each cell, and asterisks have been used to denote significance. * for $p < 0.05$, ** for $p < 0.01$, and *** for $p < 0.001$.

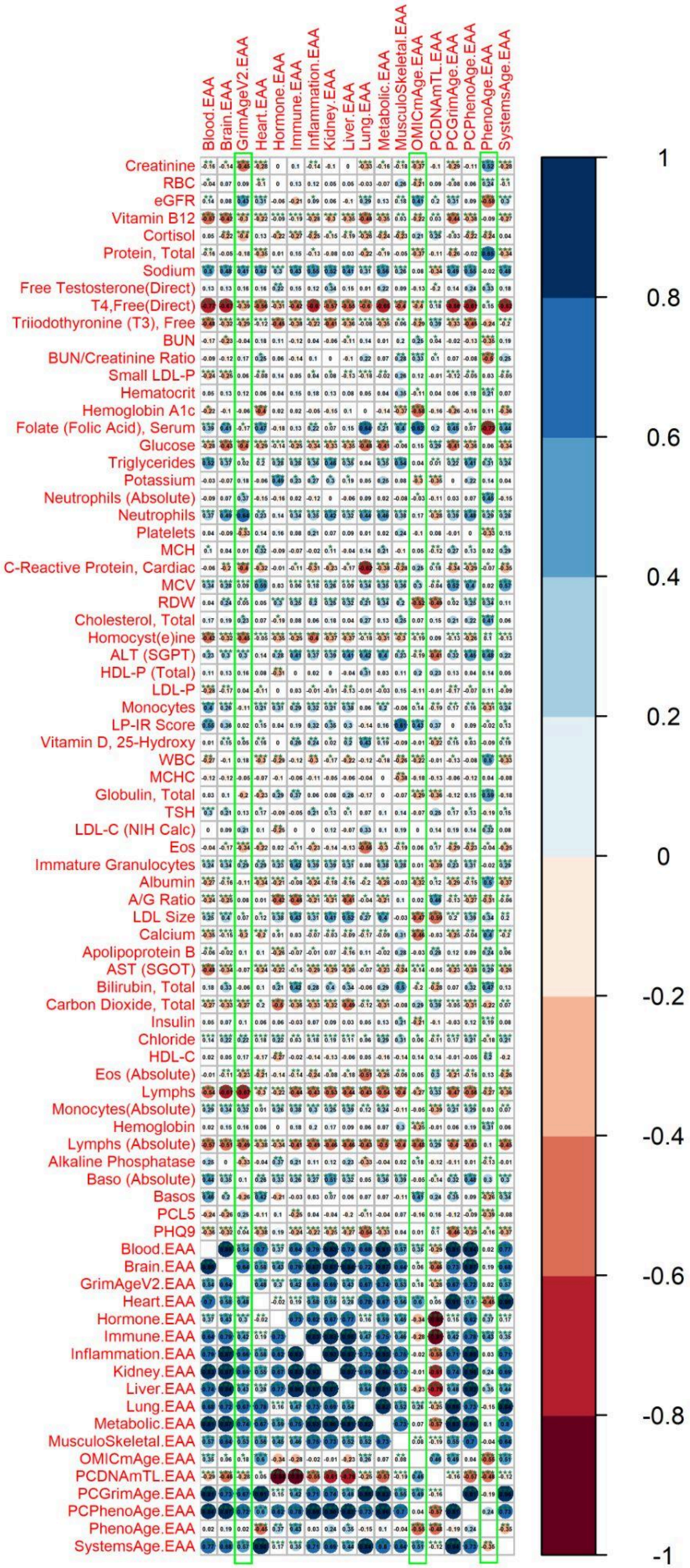

Blood.EAA
Brain.EAA
GrimAgeV2.EAA
Heart.EAA
Hormone.EAA
Immune.EAA
Inflammation.EAA
Kidney.EAA
Liver.EAA
Lung.EAA
Metabolic.EAA
MusculoSkeletal.EAA
OMICmAge.EAA
PCDNAmTL.EAA
PCGrimAge.EAA
PCPhenoAge.EAA
PhenoAge.EAA
SystemsAge.EAA
Creatinine
RBC
eGFR
Vitamin B12
Cortisol
Protein, Total
Sodium
Free Testosterone(Direct)
T4,Free(Direct)
Triiodothyronine (T3), Free
BUN
BUN/Creatinine Ratio
Small LDL-P
Hematocrit
Hemoglobin A1c
Folate (Folic Acid), Serum
Glucose
Triglycerides
Potassium
Neutrophils (Absolute)
Neutrophils
Platelets
MCH
C-Reactive Protein, Cardiac
MCV
RDW
Cholesterol, Total
Homocyst(e)ine
ALT (SGPT)
HDL-P (Total)
LDL-P
Monocytes
LP-IR Score
Vitamin D, 25-Hydroxy
WBC
MCHC
Globulin, Total
TSH
LDL-C (NIH Calc)
Eos
Immature Granulocytes
Albumin
A/G Ratio
LDL Size
Calcium
Apolipoprotein B
AST (SGOT)
Bilirubin, Total
Carbon Dioxide, Total
Insulin
Chloride
HDL-C
Eos (Absolute)
Lymphs
Monocytes(Absolute)
Hemoglobin
Lymphs (Absolute)
Alkaline Phosphatase
Baso (Absolute)
Basos
PCL5
PHQ9
Blood.EAA
Brain.EAA
GrimAgeV2.EAA
Heart.EAA
Hormone.EAA
Immune.EAA
Inflammation.EAA
Kidney.EAA
Liver.EAA
Lung.EAA
Metabolic.EAA
MusculoSkeletal.EAA
OMICmAge.EAA
PCDNAmTL.EAA
PCGrimAge.EAA
PCPhenoAge.EAA
PhenoAge.EAA
SystemsAge.EAA
1
0.8
0.6
0.4
0.2
0
-0.2
-0.4
-0.6
-0.8
-1
